# Mortality Associated with Proportionality of Secondary Mitral Regurgitation After Transcatheter Mitral Valve Repair: MFIRE Registry

**DOI:** 10.1101/2023.03.16.23287387

**Authors:** Neal M Duggal, Milo Engoren, Scott M Chadderdon, Evelio Rodriguez, M Andrew Morse, Mani A Vannan, Pradeep K Yadav, Michael Morcos, Flora Li, Mark Reisman, Enrique Garcia-Sayan, Deepa Raghunathan, Nishtha Sodhi, Paul Sorajja, Lily Chen, Jason H Rogers, Marcella A Calfon Press, Christopher P Kovach, Edward A Gill, Firas E Zahr, Stanley J Chetcuti, Yuan Yuan, Graciela B Mentz, D Scott Lim, Gorav Ailawadi

**Affiliations:** Department of Anesthesiology, University of Michigan, Ann Arbor, MI; Division of Cardiology, Knight Cardiovascular Institute, Oregon Health and Science University, Portland, OR; Department of Cardiothoracic Surgery, Ascension Saint Thomas Heart, Nashville, TN; Division of Cardiology, Ascension Saint Thomas Heart, Nashville, TN; Marcus Heart Valve Center, Piedmont Heart Institute, Atlanta, GA; Division of Cardiology, University of Washington, Seattle, WA; Department of Anesthesiology, University of Washington, Seattle, WA; Division of Cardiology, Weill Cornell Medical Center, NY, NY; Division of Cardiovascular Medicine, University of Texas Health Science Center at Houston, Houston, TX; Division of Cardiovascular Medicine, University of Virginia, Charlottesville, VA; Valve Science Center, Minneapolis Heart Institute, Abbott Northwestern Medical Center, Minneapolis, MN; Division of Cardiovascular Medicine, University of California Davis Medical Center, Sacramento, CA; Division of Cardiology, University of California Los Angeles, Los Angeles, CA; Division of Cardiology, University of Colorado, Aurora, CO; Division of Cardiovascular Medicine, University of Michigan, Ann Arbor, MI; Department of Cardiac Surgery, University of Michigan, Ann Arbor, MI

## Abstract

**BACKGROUND:** The association, if any, between the effective regurgitant orifice (EROA) to left ventricular end-diastolic volume (LVEDV) ratio and 1-year mortality is controversial in patients undergoing mitral transcatheter edge-to-edge repair (m-TEER) with the MitraClip™ system. The objective is to determine the association between EROA/LVEDV and 1-year mortality among patients undergoing m-TEER with MitraClip™.

**METHODS:** In patients with severe secondary (functional) mitral regurgitation (MR), we analyzed registry data from 11 centers using generalized linear models with the generalized estimating equations approach.

**RESULTS:** We studied 525 patients with secondary MR who underwent m-TEER in 11 centers. Most patients were male (63%) and were NYHA class III (61%) or IV (21%). MR was caused by ischemic cardiomyopathy in 51% of patients. EROA/LVEDV values varied widely with median=0.19 mm^2^/mL, interquartile range [0.12,0.28] mm^2^/mL, and 187 (36%) patients had values <0.15 mm^2^/mL. Post-procedural MR severity improved substantially, being 1+ or less in 74%, 2+ in 20%, 3+ in 4%, and 4+ in 2%. 1-year mortality was 22%. After adjustment for confounders, the logarithmic transformation of EROA/LVEDV was associated with 1-year mortality (odds ratio=0.600, 95% confidence interval [0.386,0.933], p=0.023). A higher Society of Thoracic Surgeons risk score was also associated with increased mortality.

**CONCLUSIONS:** Lower values of Ln(EROA/LVEDV) were associated with increased 1-year mortality in a multi-center registry. The slope of the association is steep at low values, but gradually becomes flatter as Ln(EROA/LVEDV) increases.

**What is Known:** In recent years, mitral transcatheter edge-to-edge repair (m-TEER) approval has expanded to select patients with functional mitral regurgitation.

The paradigm of proportionate and disproportionate MR as measured by EROA/LVEDV has been proposed as a potential rationale to reconcile conflicting trial results.

**What the Study Adds:** Before adjustment, EROA/LVEDV was not associated with mortality (0.22 ± 0.13 vs 0.21 ± 0.14, p=0.425, standardized difference = -0.08).

However, after adjustment we found that the logarithmic transformation of the pre-procedural EROA/LVEDV, Ln(EROA/LVEDV) was associated with 1-year mortality (OR = 0.600, 95% CI [0.386, 0.933], p=0.023).

Progressively higher values of pre-procedural EROA/LVEDV were always associated with a progressively lower odds ratio of death post-procedurally, without a plateau or cutoff but with an “elbow” around 0.15mm^2^/mL.

**Graphic Abstract:** The Association of EROA/LVEDV with Mortality. Plot of the adjusted odds ratios associated (solid line) with 95% confidence intervals (dashed lines) with EROA/LVEDV=0.15 as the reference showing the risk of death increases as EROA/LVEDV decreases. The plot is back transformed from Ln(EROA/LVEDV).

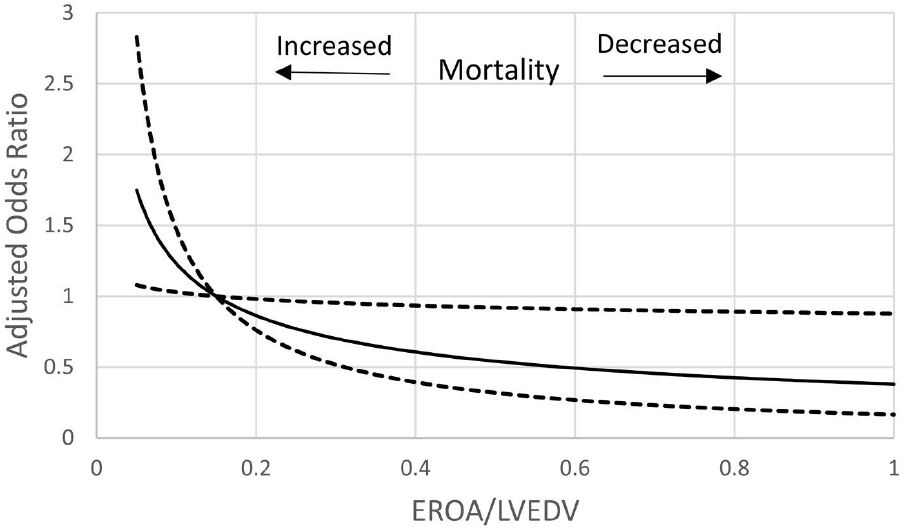

## INTRODUCTION

Unlike primary mitral regurgitation (MR), surgical intervention for secondary MR (also known as functional MR) has been shown to increase the quality of life but not extend survival.^1,2^ In these challenging patients transcatheter repair techniques have been explored as alternative treatment options and are preferred in appropriately selected patients. The largest transcatheter experience has been with the mitral transcatheter edge-to-edge leaflet repair (m-TEER) technique with the commercially-available MitraClip® device. In 2013, the FDA approved MitraClip for prohibitive surgical risk patients with primary MR based on the results of the EVEREST Trials and REALISM Registry.^3-6^ In 2019, this m-TEER approval was extended to selected patients with significant secondary MR who remain symptomatic despite maximally tolerated guideline-directed medical therapy.^6^

The FDA approval for secondary MR treatment with MitraClip integrates the results of two randomized landmark trials, COAPT (Cardiovascular Outcomes Assessment of the MitraClip Percutaneous Therapy for Heart Failure Patients with Functional Mitral Regurgitation) and MITRA-FR (Percutaneous Repair with the MitraClip Device for Severe Functional/Secondary Mitral Regurgitation).^1,2^ However, these trials have shown conflicting results of mitral valve m-TEER in patients with secondary MR and reduced ejection fraction.

The MITRA-FR study did not demonstrate a reduction of the primary combined endpoint of all-cause death and unplanned hospitalization for heart failure at 1-year follow-up.^2^ In contrast, the COAPT trial demonstrated significant reductions hospitalizations for heart failure and all-cause mortality over a 2-year follow-up.^1^

The paradigm of proportionate and disproportionate MR has been proposed as a potential rationale to reconcile these conflicting trial results.^7-9^ This concept suggests that patients enrolled in the MITRA-FR trial had MR severity that was predominantly proportional to the degree of LV enlargement (i.e. “proportionate MR”) with a smaller EROA and larger LVEDV leading to a reduced EROA/LVEDV ratio.^7-9^ In contrast, COAPT had enrolled patients whose effective regurgitant orifice (EROA) was larger but whose LV end-diastolic volumes (LVEDV) were smaller than those in MITRA-FR leading to a higher EROA/LVEDV ratio.^7-9^ Accordingly, the severity of MR was greater than could be accounted for by the degree of LV dilation (i.e. “disproportionate MR”, defined by these authors as an EROA/LVEDV>0.14 mm2/mL).^7-9^ However, questions have been raised about the volume measurements in the COAPT trial, suggesting an underestimation of LVEDV or an overestimation of the EROA.^10,11^ In addition, this model lacks enough patient-level data.^12^ More recently, registry-based studies have failed to show a consistent association between EROA/LVEDV and mortality.^13-15^

Here, we describe the association between MR proportionality, as quantified by the continuous EROA/LVEDV ratio, and mortality among patients with secondary MR and LVEF ≤55% undergoing m-TEER in the North American Mitraclip for Functional Mitral Regurgitation (M-FIRE) registry.

## METHODS

This is a multi-center retrospective study. This study received a certificate of exemption from the University of Michigan Institutional Review Board (HUM00191906, Ann Arbor, MI). As no patient care interventions were involved, signed patient consent was waived. N Duggal had full access to all the data in the study and takes responsibility for its integrity and the data analysis. We included registry data from 11 MitraClip centers (Appendix 1). We included all adult patients (≥18 years) with symptomatic (New York Heart Association II, III, IV) MR assessed by echocardiography within 3-6 months of undergoing m-TEER with the MitraClip™ device, and left ventricular ejection fraction ≤55%.^16^ Patients were excluded if they had concomitant primary/degenerative MR (prolapse/flail leaflets), secondary MR predominantly driven by mitral annular dilatation (atrial FMR) with presence of normal ventricular function, prior mitral valve leaflet surgery, cardiogenic shock requiring need for mechanical support preoperatively, status 1 heart transplant listed or previous heart transplant, or active endocarditis or evidence of intracardiac thrombus.

All studies were interpreted by expert echocardiographers at large, tertiary care institutions, and the severity of MR before and after the intervention was determined based on the multi-parametric approach and quantitation methods recommended by the American Society of Echocardiography guidelines.^16-18^ MR was classified using a 4-grade system as recommended by the Mitral Valve Academic Research Consortium: mild (1+), moderate (2+), moderate-severe (3+), or severe (4+).^19^ Left ventricular internal dimensions in systole and diastole were obtained via the parasternal long-axis view in transthoracic echocardiograms.. The left ventricular volumetric measurements were obtained using two-dimensional Simpson’s method of discs.^17^ The follow up studies were transthoracic studies and assessment of MR was assessed by post-procedure valvular regurgitation guidelines.^18^

### DATA COLLECTION

Study data was collected via combined queries from each participating institution’s electronic medical record and the Society of Thoracic Surgeons (STS) & American College of Cardiology (ACC) Transcatheter Valve Therapy (TVT) Registry from 2014 to 2020. They were then combined into the M-FIRE registry. Within the STS/ACC TVT registry database patient history, demographics, and preoperative echocardiographic, MitraClip procedural, postoperative, and follow-up data are stored as discrete, searchable database concepts. Echocardiographic data were reviewed by each of the participating sites.

### STATISTICAL ANALYSIS

For the outcome of 1-year mortality, patients who were alive at 1 year were compared to those who died by 1 year using Fisher’s exact test for categorical variables, ANOVA F-test for continuous variables, and standardized differences. P values <0.05 were deemed statistically significant and standardized differences >0.20 or <-0.20 were deemed clinically important. Continuous variables are expressed as mean ± standard deviation or median [interquartile range]. Next, generalized estimating equations (to account for the clustering within each center) were used to assess associations between EROA/LVEDV and 1-year mortality. Models were adjusted for patient characteristics, STS risk score, and other echocardiographic factors. Coefficients are expressed as odds ratio (OR) and 95% confidence intervals [95% CI]. Continuous variables were assessed for linearity and if nonlinear were transformed as appropriate. Echocardiographic measurements are frequently log-transformed to decrease skew, decrease heteroscedasticity, and improve model fit.^20,21^ Variables were assessed for collinearity using variance inflation factor >5 and for correlation using Pearson correlation coefficient >0.7.

Missing demographic and cardiac parameters were imputed with chained equations and combined using Rubin’s Rules. Missing mortality data were handled in a sensitivity analysis by imputing the outcomes with chained equations. Generalized linear model with the generalized estimating equations approach were conducted as above, with exchangeable correlation matrix and logit link. 95% confidence intervals of the odds ratios that excluded 1 were deemed statistically significant. All statistics were conducted in SAS (SAS Institute, Cary, NC) and SPSS 27 (IBM, Chicago, IL).

### POWER ANALYSIS

Assuming 20% mortality, an intracluster correlation = 0.3, and 10 participating centers, having 346-379 patients would give us 80% power with alpha = 0.05 to find a change in mortality to 18% for a 20% increase in the EROA/LVEDV ratio and to have 5 predictors in the regression.

## RESULTS

### STUDY POPULATION AND BASELINE CHARACTERISTICS

In eleven American tertiary care heart valve centers, 525 patients were treated with m-TEER for secondary MR between 2014 and 2020 and were included in the M-FIRE registry. After excluding 33 patients (6%) with unknown mortality status, the primary analysis consisted of 492 patients. At one-year follow-up, 108 (22%) had died and 384 (78%) were alive. The baseline characteristics of the patients who were alive or dead at one-year post-procedure are summarized in Tables 1 and 2. The majority of patients were male (63%), with a history of hypertension (84%). Ischemic cardiomyopathy was the most common etiology of MR (51% of patients). Most patients were NYHA functional class III and IV, 61% and 21%, respectively. Pre-procedural MR severity was 3+ in 122 patients (23%) and 4+ in 383 patients (73%) (Table 3). EROA/LVEDV values varied widely with median=0.19, interquartile range [0.12,0.28] mm^2^/mL, and 187 (36%) patients had values <0.15 mm^2^/mL(Figure 1). Post-procedural MR severity was much improved, being 1+ or less in 367 patients (74%), 2+ in 99 patients (20%), 3+ in 16 patients (3%), and 4+ in 10 patients (2%).

**Table 1.** Patient Characteristics – Categorical Factors

| Factor | All<br>(n=525) |  | Alive<br>(n=384) |  | Died<br>(n=108) |  | StandDiff | P-value |
| --- | --- | --- | --- | --- | --- | --- | --- | --- |
|  | n | % | n | % | n | % |  |  |
| Male | 328 | 63 | 236 | 61 | 73 | 68 | -0.13 | 0.261 |
| Diabetes mellitus | 182 | 35 | 132 | 34 | 38 | 35 | 0.02 | 0.909 |
| Hypertension | 439 | 84 | 333 | 87 | 81 | 75 | -0.28 | 0.010 |
| Previous myocardial infarction | 250 | 48 | 182 | 47 | 54 | 50 | 0.05 | 0.664 |
| Previous percutaneous coronary intervention | 202 | 39 | 149 | 39 | 42 | 39 | 0.00 | 0.999 |
| Previous coronary artery bypass grafting | 171 | 33 | 120 | 31 | 41 | 38 | 0.14 | 0.202 |
| Previous stroke | 74 | 14 | 59 | 15 | 14 | 13 | -0.07 | 0.646 |
| COPD | 195 | 37 | 138 | 36 | 48 | 44 | 0.17 | 0.118 |
| History of atrial fibrillation or atrial flutter | 313 | 60 | 231 | 60 | 68 | 63 | 0.06 | 0.656 |
| Ischemic mitral regurgitation* | 269 | 51 | 186 | 48 | 66 | 61 | 0.26 | 0.022 |
| Pacemaker/Cardiac resynchronization therapy | 192 | 37 | 138 | 36 | 41 | 41 | 0.12 | 0.311 |
| Internal defibrillator | 238 | 45 | 174 | 46 | 49 | 49 | 0.05 | 0.662 |
| New York Heart Association Class |  |  |  |  |  |  | 0.25 | 0.278 |
| I | 6 | 1 | 6 | 2 | 0 | 0 |  |  |
| II | 89 | 17 | 65 | 17 | 12 | 11 |  |  |
| III | 319 | 61 | 231 | 61 | 69 | 64 |  |  |
| IV | 111 | 21 | 78 | 21 | 26 | 24 |  |  |

Thirty-three patients are missing 1 year outcome. Data are missing for 1 patient for sex, diabetes mellitus, hypertension, previous percutaneous coronary intervention, previous coronary artery bypass grafting, previous stroke, and history of atrial fibrillation or atrial flutter, 4 for previous myocardial infarction and ischemic mitral regurgitation, 6 for New York Heart Association class, and 9 for COPD. \*included 2 with mixed ischemic/non-ischemic etiology. COPD-chronic obstructive pulmonary disease. StandDiff-standardized difference.

**Table 2.** Patient Characteristics -Continuous Factors.

| Factor | All<br>(N=525) |  | Alive<br>(n=384) |  | Died<br>(n=108) |  | StandDiff | P-value |
| --- | --- | --- | --- | --- | --- | --- | --- | --- |
|  | Mean | SD | Mean | SD | Mean | SD |  |  |
| Age (yr) | 71 | 13 | 71 | 12 | 72 | 13 | 0.14 | 0.202 |
| Height (cm) | 171 | 11 | 171 | 11 | 172 | 9 | 0.15 | 0.184 |
| Weight (kg) | 79 | 21 | 80 | 21 | 78 | 18 | -0.07 | 0.534 |
| Body Mass Index (kg/m<sup>2</sup>) | 27.0 | 6.0 | 27.2 | 6.1 | 26.3 | 5.5 | -0.16 | 0.154 |
| Creatinine (mg/dL) | 1.7 | 1.5 | 1.7 | 1.9 | 2.1 | 1.9 | 0.29 | 0.004 |
| STS Replacement risk (%) | 8.8 | 7.8 | 7.4 | 6.4 | 12.6 | 10.1 | 0.61 | <0.001 |

Thirty-three patients are missing 1 year outcome. Data are missing for 3 patients for age, 8 for height, 1 weight, 1 creatinine, and 47 STS replacement risk score. SD-standard deviation. StandDiff-standardized difference. STS Replacement-Society of Thoracic Surgeons risk estimate of mortality with a mitral valve replacement.

**Table 3.** Patient Echocardiography Measurements

| Factor | All<br>(n=525) |  | Alive<br>(n=384) |  | Died<br>(n=108) |  |  |  |
| --- | --- | --- | --- | --- | --- | --- | --- | --- |
|  | n | % | n | % | n | % | StandDiff | P-value |
| Mitral regurgitation severity |  |  |  |  |  |  | 0.14 | 0.595 |
| I | 1 | 0.2 | 1 | 0.3 | 0 | 0 |  |  |
| II | 19 | 4 | 12 | 3 | 6 | 6 |  |  |
| III | 122 | 23 | 90 | 23 | 25 | 23 |  |  |
| IV | 383 | 73 | 281 | 73 | 77 | 71 |  |  |
|  | Mean | SD | Mean | SD | Mean | SD | StandDiff | P-value |
| LVEDV (mL) | 201 | 84 | 199 | 81 | 209 | 89 | 0.12 | 0.238 |
| EROA (cm<sup>2</sup>) | 0.38 | 0.18 | 0.38 | 0.19 | 0.38 | 0.16 | -0.08 | 0.501 |
| EROA/LVEDV (mm<sup>2</sup>/mL) | 0.22 | 0.15 | 0.22 | .13 | 0.21 | 0.14 | -0.09 | 0.425 |
| Ln(EROA/LVEDV) | -1.68 | 0.59 | -1.66 | 0.57 | -1.74 | 0.59 | -0.13 | 0.223 |
| LVESV (mL) | 139 | 69 | 136 | 67 | 147 | 73 | 0.16 | 0.138 |
| Ejection fraction (%) | 0.32 | 0.10 | 0.33 | .10 | 0.31 | .10 | -0.27 | 0.040 |
| Discharge MV gradient (mmHg) | 4 | 2 | 4 | 2 | 5 | 2 | 0.15 | 0.179 |
|  | Median | IQR | Median | IQR | Median | IQR | StandDiff | P-value |
| EROA/LVEDV (mm<sup>2</sup>/mL) | 0.19 | 0.12, 0.28 | 0.19 | 0.13, 0.28 | 0.17 | 0.11, 0.24 | -0.08 | 0.107 |
| Ln(EROA/LVEDV) | -1.68 | -2.10, -1.29 | -1.66 | -2.00, -1.28 | -1.76 | -2.17, -1.42 | -0.09 | 0.107 |
| TR severity | 2 | 1, 3 | 2 | 1, 3 | 3 | 1, 3 | 0.31 | 0.136 |
| RV dysfunction | 1 | 0, 2 | 1 | 0, 2 | 1 | 0.5, 2 | 0.39 | 0.007 |
| Post MR severity | 1 | 1, 1.5 | 1 | 1, 1 | 1 | 1, 2 | 0.50 | 0.001 |
| Post MV gradient (mmHg) | 3 | 2, 4 | 3 | 2, 4 | 3 | 2, 5 | 0.23 | 0.049 |
| Discharge MR severity | 1 | 1, 2 | 1 | 1, 1.5 | 1 | 1, 2 | 0.41 | 0.001 |

Data are missing for 18 patients for discharge MV gradient, 8 RV dysfunction, 1 post MR severity, and 14 discharge MR severity. EROA-estimated regurgitant orifice area. IQR-interquartile range. Ln(EROA/LVEDV)-natural logarithm of estimated regurgitant orifice area/left ventricular end-diastolic volume. LVEDV-left ventricular end-diastolic volume. LVESV-left ventricular end-systolic
volume. MR-mitral valve regurgitation. MV-mitral valve. RV-right ventricle. SD-standard deviation. StandDiff-standardized difference. TR-tricuspid valve regurgitation.

**Figure 1.**
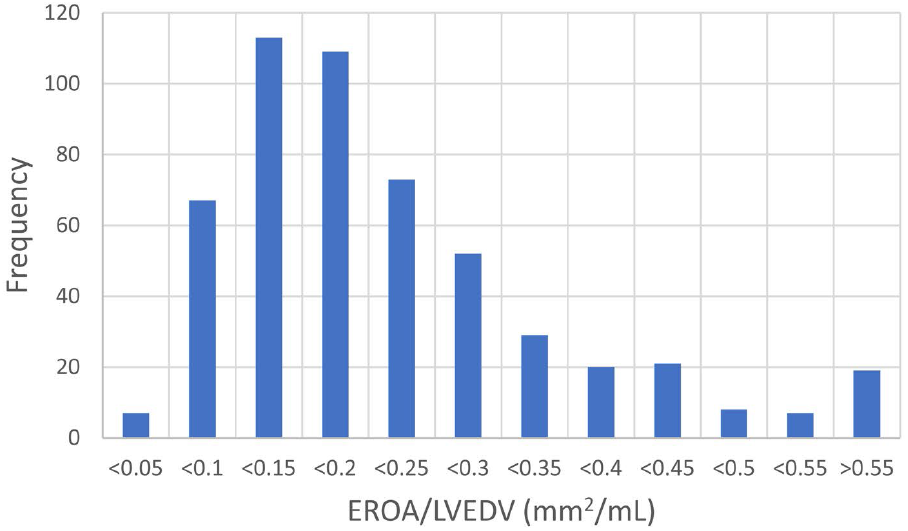
Histogram showing the frequencies of the effective regurgitant orifice area divided by the left ventricular end diastolic volume (EROA/LVEDV) sizes. Median=0.19 and interquartile range [0.12, 0.28] mm^2^/mL. 187 (36%) patients had values <0.15 mm^2^/mL

### MORTALITY

There were no differences in patient demographics between those who were alive or dead at one year, however, STS replacement risk (12.6% ± 10.1 vs 7.4% ± 6.4, p≤0.001) was higher in the patients who died (Table 2). Univariate analysis revealed that several pre-procedural echocardiographic parameters, such as ejection fraction and right ventricular dysfunction, were significantly associated with 1-year mortality. Notably, EROA/LVEDV was not associated with mortality (0.22 ± 0.13 vs 0.21 ± 0.14, p=0.425, standardized difference = -0.08) (Table 3). Post-procedure, worse MR was associated with increased mortality 1+ [1+,2+] vs 1+[1+,1+], p=0.001, standardized difference=0.50) (Table 3).

Using generalized estimating equations to adjust for confounders, we found that the logarithmic transformation of the pre-procedural EROA/LVEDV, Ln(EROA/LVEDV), was associated with 1-year mortality (OR=0.600, 95% CI [0.386, 0.933], p=0.023) (Table 3). On back-transformation, the odds of dying initially decreased rapidly, but then gradually became less steep. Progressively higher values of pre-procedural EROA/LVEDV were always associated with a progressively lower odds ratio of death post-procedurally, without a plateau or cutoff but with an “elbow” around 0.15mm^2^/mL. Using EROA/LVEDV=0.15 mm^2^/mL having an OR = 1.0 as the reference, an EROA/LVEDV=0.1 mm^2^/mL is associated with an 23% [3%, 47%], p<0.001 greater odds of dying (OR=1.23, 95% CI [1.03, 1.47]). At EROA/LVEDV values greater than 0.15 mm^2^/mL, the OR for mortality is < 1 and decreases more slowly, being 0.70 [0.52,0.95], p<0.001 at an EROA/LVEDV=0.3 mm^2^/mL and 0.49 [0.27, 0.91], p<0.001 at EROA/LVEDV=0.6 mm^2^/mL (Graphic Abstract). We also found that having a higher STS replacement risk score (OR=1.09, 95% CI [1.05, 1.13], p≤0.001), was also independently associated with increased mortality (Table 4).

**Table 4.** Factors Associated with 1 Year Mortality

| Factor | Odds ratio | 95% Confidence Interval | P-value |
| --- | --- | --- | --- |
| Ln(EROA/LVEDV) | 0.600 | 0.386,0.933 | 0.023 |
| Hypertension | 0.312 | 0.174,0.559 | <0.001 |
| STS replacement % | 1.090 | 1.053,1.128 | <0.001 |
| COPD | 1.517 | 0.945,2.430 | 0.084 |
| Age (yr) | 1.003 | 0.982,1.024 | 0.789 |
| Body mass index (kg/m<sup>2</sup>) | 0.987 | 0.946,1.030 | 0.548 |
| Female | 0.780 | 0.481,1.265 | 0.314 |
| NYHA class 3 | 1.744 | 0.861,3.534 | 0.123 |
| NYHA class 4 | 1.384 | 0.620,3.091 | 0.427 |
| Intercept | 0.093 | 0.011,0.804 | 0.031 |

Parsimonious Logistic regression showing the factors independently associated with death at 1 year follow-up. COPD-chronic obstructive pulmonary disease. Ln(EROA/LVEDV)-natural logarithm of estimated regurgitant orifice area/left ventricular end-diastolic volume. NYHA-New York Heart Association. STS-Society of Thoracic Surgeons.

### SENSITIVITY ANALYSIS

To try to determine the possible effect of the 33 (6%) patients missing survival outcomes on our findings, we performed a sensitivity analysis by using multiple imputation. In this analysis we found Ln(EROA/LVEDV) was still associated with 1-year mortality (OR=0.62, 95% CI [0.40, 0.97], p=0.035.

## DISCUSSION

We found that after adjusting for patient characteristics, STS predicted replacement risk and the echocardiographic measurement of Ln(EROA/LVEDV) was associated with mortality. We found that the association was very steep (Graphic Abstract) at lower values of EROA/LVEDV, but became flatter at higher values, suggestive of an elbow at ∼0.15 mm^2^/mL but no plateau or cutoff. Our findings suggest that despite a good repair, as measured by residual MR and mitral gradient, the pre-m-TEER pathology with patients having lower EROA/LVEDV values have long-lasting effects on mortality. Study is needed to determine how best to treat these high-risk patients. Our findings of an elbow at ∼0.15 mm^2^/mL also suggest that the mechanisms associated with MR and m-TEER mortality may differ consistent with the proportionality theory.

Two randomized controlled trials published in 2018 examining m-TEER in patients with chronic heart failure and secondary MR noted conflicting results. In the MITRA-FR trial m-TEER did not reduce the risk of death or hospitalization for heart failure at 2 year follow up. On the contrary, COAPT trial led to significant reduction in mortality and hospitalization for heart failure over the 2 year follow up.^1,2^ Numerous hypotheses have been discussed as plausible explanations for the discordant results of these two trials. In addition to noting the different definitions of severe secondary MR and differences in baseline LV volumes between these trials, there has been much interest in determining whether a severity of MR that is disproportionate to the degree of LV dilatation could predict clinical outcomes. Grayburn et al. developed a pathophysiologic premise that the two trials enrolled different patient populations and that EROA/LVEDV ratio could be useful in clinical decision-making.^7^ They noted in MITRA-FR that MR severity could be explained by the degree of LV enlargement, i.e., proportionate MR with low EROA/LVEDV which would likely respond to optimization of medical therapy. Whereas, in COAPT, MR severity was much greater than could be accounted for by degree of LV enlargement, i.e., disproportionate MR with high EROA/LVEDV ratios and likely to benefit from m-TEER. These authors designated proportionate MR by an EROA/LVEDV of “roughly <0.14 to 0.15 mm^2^/mL” and proposed this metric to help differentiate between disproportionate and proportionate MR.^7^

Several studies investigating this hypothesis, however, have demonstrated limitations on its ability to help identify those patients most likely to benefit from m-TEER. Orban et al. divided the patients in a large multicenter European registry (EuroSMR) of over 1000 patients into tertiles of EROA/LVEDV (<0.115, between 0.115 and 0.165, and ≥0.165 mm^2^/mL) and found only a small increased 2-year mortality in the group with EROA/LVEDV <0.115 mm^2^/mL but no difference between the other 2 groups^13^ In a multicenter registry study, Ooms et al. investigated several EROA/LVEDV cutoffs before settling on the median value 0.13 mm^2^/mL.^14^. They found no difference in 2-year mortality between the 2 groups. Similarly, Adamo et al. in a smaller retrospective multicenter registry noted no independent prognostic value in EROA/LVEDV, when dichotomized at the median value 2.25 cm^2^/L, which is equal to 0.225 mm^2^/mL.^15^ Furthermore, post hoc analysis of MITRA-FR noted neutral impact of m-TEER above and below numerous EROA/LVEDV ratios.^22^ Lastly, post hoc analysis of COAPT failed to demonstrate that EROA/LVEDV appropriately identified responders to m-TEER.^23^ Our study differs from these studies by analyzing EROA/LVEDV as a continuous variable, rather than dichotomizing it. Rarely are there abrupt or dichotomous outcomes from a physiologic parameter. Instead, the probability of the outcome varies smoothly as the physiologic parameter changes. Dichotomizing EROA/LVEDV creates the appearance of an abrupt change at the cutoff, loses information and statistical power (equivalent to losing more than one-third of the patients), and creates false dichotomies, such that all values on the same side of the cutoff have the same risk.^24-26^ Instead, we found that the risk associated with EROA/LVEDV continuously varies, but that the rate at which risk varies depends on the EROA/LVEDV value. At low values of EROA/LVEDV, the change in risk for a given change in EROA/LVEDV is larger than it is at a larger EROA/LVEDV. Hence small differences in measurement of proximal isovelocity surface area (PISA) to estimate EROA will lead to larger changes in estimated risk at low values of EROA/LVEDV than at higher values of EROA/LVEDV.

### STUDY LIMITATIONS

First, this is an observational, non-randomized, and retrospective study. As such, we acknowledge the limitations in reporting as well as selection bias. However, the study was conducted in a real world setting where the evaluation and indication for m-TEER was determined by expert heart teams at large experienced tertiary care centers. In addition, all patients were deemed high or prohibitive surgical risk by the local heart team (in contrast to COAPT). With the absence of an independent echocardiography core laboratory, interobserver variability cannot be excluded. However, all measurements were performed by experienced echocardiographers according to the current American Society of Echocardiography guidelines and recommendations.^16-18^ In addition, as 3D assessment has better correlation with cardiac MRI, it is possible that 2D LV size and EROA may have been underestimated. Another limitation is that 6% of patients were missing 1-year outcomes. We conducted a sensitivity analysis using imputation and found that the association between Ln(EROA/LVEDV) and 1 year mortality was robust to these missing 33 patients.

### STUDY STRENGTHS

The multicenter basis should improve the generalizability of our results. Importantly, rather than just defaulting to a linear relationship between EROA/LVEDV and mortality, we assessed for the shape of the association, which was best with a logarithmic transformation. This is similar to other studies of echocardiographic measurements that also used logarithmic transformations.^20,21^

## CONCLUSIONS

Using the M-FIRE registry, we demonstrated that proportionality, as defined by EROA/LVEDV, provides prognostication of mortality risk after m-TEER. We found that the association was very steep (Graphic Abstract) at lower values of EROA/LVEDV, became flatter at higher values, but without a plateau or cutoff. Our findings contribute to the ongoing debate while providing new insights. Equally, our findings call for further investigation in prospective studies. In conclusion, we found that lower values of Ln(EROA/LVEDV) are associated with higher 1 year mortality.

## Data Availability

Study data was collected via combined queries from each participating institution's electronic medical record and the Society of Thoracic Surgeons (STS) & American College of Cardiology (ACC) Transcatheter Valve Therapy (TVT) Registry. Echocardiographic data were reviewed by each of the participating sites.

## Acknowledgments

No additional acknowledgements.

## Abbreviations

CI: confidence interval
COAPT: Cardiovascular Outcomes Assessment of the MitraClip Percutaneous Therapy for Heart Failure Patients with Functional Mitral Regurgitation
EROA: effective regurgitant orifice area
FMR: functional mitral regurgitation
LVEDV: left ventricular end diastolic volume
LVEF: left ventricle ejection fraction
M-FIRE: North American Mitraclip for Functional Mitral Regurgitation
MR: mitral regurgitation
m-TEER: mitral transcatheter edge to edge repair
OR: odds ratio
TVT: Transcatheter Valve Therapy registry

